# Parental experiences of the impacts of COVID-19 on the care of young children; qualitative interview findings from the Nairobi Early Childcare in Slums (NECS) Project

**DOI:** 10.1101/2022.09.09.22279760

**Authors:** Robert C Hughes, Ruth Muendo, Sunil S Bhopal, Silas Onyango, Elizabeth Kimani-Murage, Betty R Kirkwood, Zelee Hill, Patricia Kitsao-Wekulo

**Affiliations:** Department of Population Health, Faculty of Epidemiology & Population Health, London School of Hygiene & Tropical Medicine, Keppel Street, London WC1E 7HT; Human Development Theme, African Population and Health Research Center, Nairobi, Kenya; Population Health Sciences Institute, Faculty of Medical Sciences, Newcastle University, Newcastle upon Tyne, Tyne and Wear, UK; Health and Wellbeing Theme, African Population and Health Research Center, Nairobi, Kenya; Institute for Global Health, University College London, London, UK

**Keywords:** Covid-19, Early Childhood Development, Urban health, Child health, Childcare, Nurturing Care

## Abstract

**Introduction:** The Covid-19 pandemic, and societal attempts to control it, have touched almost every aspect of people’s lives around the world, albeit in unequal ways. In particular, there is considerable concern about the way that stringent ‘lockdowns’, as implemented in Kenya and many other countries, affected young children, especially those living in informal settlements. However, to date, there has been little research attempting to unpack and understand how the pandemic has impacted on the care of young children.

**Methods:** In-depth telephone interviews were conducted with 21 parents/carers of children aged under five years living in three Nairobi slums between May and September 2021 exploring the ways in which covid-19, and policies to control the pandemic, impacted on their household and the care of their child/children.

**Results:** The impacts of covid-19 control measures on the care of children have been widely felt, deep and multiple. The impact of economic hardship has been significant, reportedly undermining food security and access to services including healthcare and childcare. Respondents reported an associated increase in domestic and community violence. Many people relied on help from others; this was most commonly reported to be in the form of variable levels of flexibility from landlords and help from other community members. No direct harms from covid-19 disease were reported by respondents.

**Conclusion:** The impacts of covid-19 control measures on the care of young children in informal settlements have been indirect but dramatic. Given the breadth and depth of these reported impacts, and the particular vulnerability of young children, deeper consideration ought to inform decisions about approaches to implementation of stringent disease control measures in future. In addition, these findings imply a need for both short- and long-term policy responses to ameliorate the impacts described.

**Key messages:** - **Young children living in slums, while at low direct risk from Covid-19, are highly vulnerable to early childhood adversity**, so may be at great risk from economic and other hardships that are a likely ‘side effect’ of blunt pandemic control measures like stringent ‘lockdowns’.
- **Parent/carers described a set of indirect impacts of covid-19 control efforts that were broad, deep and protracted**. Core to these impacts was widespread economic hardship, with knock on effects on household food security, wellbeing and community safety.
- **Considering the particular risks and vulnerability that blunt pandemic control measures present to young children, especially those in slums, needs to be central to policy discussions about if and how to implement stringent disease-control measures**. In addition, more research is required to quantify the issues identified in this qualitative inquiry.

## Introduction

The Covid-19pandemic has touched the lives of almost everyone on the planet, but in very different ways. In Kenya, there was an early and stringent response to the first cases of community transmission, including one of the most harshly enforced ‘lockdowns’ in the world(1). Efforts to control the pandemic in Nairobi were especially felt in the informal settlements where 60% of the city’s population live (2). Enforcement of lockdowns was strict, with reports of violence and heavy-handed crackdowns from police especially in informal settlements (3).

Early childhood is a critical window of opportunity; adversity in this period is a central social determinant of health and wellbeing, affecting later life learning, earning and happiness(4). Early in the pandemic, concerns were raised about how the control measures would be likely to impact on young children, anticipating that “vulnerable children will bear the biggest brunt of the direct and indirect impacts of the pandemic”. Shumba and colleagues (2020) noted that in addition to direct health impacts from Covid-19, young children are also at risk from impacts on health, nutrition, social and child protection systems alongside economic disruption (5). However, little research has been published to date attempting to explore the lived experiences of these impacts as they have emerged, especially in low- and middle-income countries. In particular, we are unaware of any other research which has sought to gain an in-depth understanding of parent’s/carers’ experiences of the impacts of Covid-19 on the care of young children living in urban slums in Africa.

We aimed to contribute to addressing this research gap through conducting in-depth telephone interviews with parents/carers from across three slums in Nairobi to gain an understanding of their experiences of caring for a child in this context at a time of covid. The results presented here are part of the larger Nairobi Early Childcare in Slums (NECS) study which through mixed-methods aims to understand the use, provision and quality of paid childcare in an informal settlement in Kenya(6).

## Methods

### Study Design

Qualitative in-depth interviews, conducted remotely by telephone.

### Timing, setting and participant characteristics

In-depth telephone interviews were conducted between 11^th^ May and 17^th^ September 2021, with parents/carers of children aged under 5 years who were living in one of three slums in Nairobi (Kibera, Kawangware and Mukuru-Viwandani). At this time, Kenya experienced a fourth wave of covid-19, with the 7-day average number of reported cases ranging between 263 and 1974(7). At the time of the interviews, covid-19 control measures were ongoing, albeit much less stringent than early in the pandemic. These consisted of night curfews (from 8pm and then later from 10pm), mask mandates in public areas, limits on public gatherings and advice to work from home(8).

The three slums were selected primarily because collectively they are typical of the larger and longer established slums across Nairobi in which the majority of the population live. Practical considerations were relevant too; this sample was drawn from an existing database of telephone numbers for low-income households who had agreed to being invited to take part in research that our data collection partner, BUSARA(9), had previously collated.

These three slums are characterised by widespread poverty, poor water and sanitation provision, inadequate shelter, insufficient infrastructure, high levels of insecurity and high rates of informal employment(10). All three slums are well established, and have existed for decades. The ethnicity across all is mixed, as is the mix of new arrivals (including rural-urban migrants and international migrants) and long-standing residents. Each slum is loosely divided into villages, which tend to be dominated by one ethnic group, but boundaries are frequently blurred. Data on employment and education enrolment in the slums are limited, but a recent study found that approximately 50% of the population in Viwandani had completed secondary or more schooling and around a third of females and 8% of males were unemployed, noting that most work in the informal sector (11).

### Data collection

RM, an experienced interviewer with Masters level training, in Development Studies, conducted telephone interviews in Kiswahili using a semi-structured topic guide developed by all authors (Supplementary Appendix 1). The content of the topic guide included both questions about covid-19 impacts alongside a broader set of themes about childcare in the slums (manuscript in preparation). This topic guide was informed by a literature review, including the review conducted by Shumba and colleagues(5) which considered how the domains of the Nurturing Care Framework (Health, Nutrition, Responsive caregiving, Early learning, and Security and Safety) would be likely to be affected by Covid-19 and/or pandemic control measures. Interview topic guides were iteratively modified as needed over the course of data collection, building on experiences, perceptions and ideas that emerged.

Selection of participants was as follows. First, a list of respondents who had completed up to 5 rounds of a bi-monthly structured telephone survey tracking the impact of Covid-19 on the care of children in slums (12) was randomly ordered. Next, RM worked through this list, selecting participants purposively, including a mixture of both male and female parents/carers of a variety of ages of children, and both users and non-users of paid childcare. When participants with specific characteristics were sufficiently well represented in the sample, potential participants on this list were skipped until a participant with a desired characteristic was reached.

Timing of telephone interviews was pre-arranged through a recruitment call. For the interview itself, participants were asked to find a quiet place to take the call. Calls started with RM introducing herself and reading participant information and consent scripts. Where necessary this information was re-phrased to explain it clearly to participants to ensure understanding and any emerging questions were answered by RM.

Interviews were digitally audio recorded. They were then simultaneously transcribed and translated verbatim into English by a professional translator. Batches of 1-3 translated transcripts were reviewed and, where needed, corrected by RM in advance of analysis. RCH, RM, PK-W, SO and ZH met approximately weekly during fieldwork to review transcripts and field notes and identify and discuss emerging themes, modifying the topic guide where necessary to allow deeper exploration of emerging important themes. In addition, these meetings were used to discuss the emergence of saturation, when it was felt that further interviews would be unlikely to lead to additional insights. No repeat interviews were carried out.

### Public involvement

Community engagement meetings were held in advance of the broader NECS Study in February 2020, introducing the study. Because this was before the Covid-19 pandemic, these did not specifically discuss the issue of pandemic impacts/controls. During preparation of this manuscript emerging findings were shared in a community meeting in Nairobi in March 2022.

### Ethical considerations

At the start of interviews, an information script was read out and participants were asked to confirm that they agreed (1) to take part, (2) for the conversation to be recorded, translated and transcribed, and (3) for these data and results to be shared and used with researchers and others both in and outside of Kenya. This verbal consent process was audio-recorded. LSHTM Research Ethics Committee (LSHTM Ref: 22692) and Amref Health Africa’s Ethics and Scientific Review Committees (ESRC) in Kenya (Ref: P777/2020) have reviewed and approved the study protocol. The National Commission for Science, Technology and Innovation (NACOSTI) provided research clearance.

### Data analysis

Data analysis was concurrent with data collection through regular weekly team discussions and a combination of iterative and deductive coding. Transcripts were read several times to build familiarity with the data and were then coded by RCH using NVivo 12(13). This started with a simple high level coding template of key headings based on domains of the Nurturing Care Framework(14), to which sub-themes or emerging themes were added.

Throughout, the focus was on understanding the underlying meaning behind statements and identifying widely held or contradictory responses/themes. Sub-themes and draft coding schedules were shared and discussed at regular intervals amongst the authors, and reflective notes were kept throughout the process. Through these discussions, the key themes presented below were identified.

The epistemological position of the researchers was discussed before and during analysis; with the team adopting a pragmatic position(15), seeking to focus on the utility of knowledge to inform policy, programmes and interventions. This research is also phenomenological, in that it seeks to draw on the experiences of research participants’ own descriptions of their lives.

RM, SO and PW are mixed methods early childhood development researchers living and working in Kenya. RCH, ZH, SB and BK are UK-based child health and development researchers. RCH has worked as a health adviser at several international donor organisations. SB is a practising community child health physician.

## Results

A total of 21 interviews were conducted. These took between 14 and 39 minutes, including the broader ranging discussion about childcare in slums but excluding the informed consent process. The mean duration was 22 minutes. All of the participants approached agreed to take part in the study.

The characteristics of the sample are described in detail in Supplementary Table 1. In summary it comprised 11 mothers, 8 fathers and 2 grandparents with similar number of users (n=11) and non-users (n=10) of paid childcare. Around half of the participants had children aged 12-23 months (n=10).

Analysis identified three key themes. Firstly, indirect impacts of covid-19 controls were more significant than direct effects of the virus. Secondly, these impacts were broad, and affected all domains of Nurturing Care. Finally, help, where it was available generally came from within the community rather than from the government.

### Indirect impacts of covid-19 controls were more significant than direct effects of covid-19

The first major theme identified was that the indirect impacts of covid-19 control measures, in particular economic hardship, were more significant than the direct effects of covid-19. The impacts of efforts to control the covid-19 pandemic on the care of children in slums were described as significant and multi-faceted by all respondents. There was a universal sense that the pandemic had affected people and their daily lives deeply. However, all of the effects were indirect; none of the interviewees described knowingly suffering from covid-19 infection themselves, or their children becoming unwell with the disease.

Economic effects were described by almost all respondents, with the loss of jobs and of informal income generating opportunities affecting those working in a variety of roles and sectors, including domestic work, factory work, market trading and informal ‘piece work’ or daily labouring. All of these became even less reliable sources of income:

> Money has reduced. There is no money [but] needs are still many. …those things, even paying the house, has become a problem…. Because there is no way you will get to pay you have to struggle… and sometimes you find you don’t get. Surviving means doing any work that you will get. – IDI15
>
> The economic impacts were described as cross-cutting, and affecting the whole community, at times leading to evictions, loss of household assets or changes in income-generating activities: People are indoors so there are no jobs. … We have hope you know. [But it is a] hard life. When the economy is down everyone is affected, we take home what we get and the costs rises… Things are not good, sometimes you will find some friends lost their jobs and sometimes they want a handout and maybe you don’t have. And sometimes you are late on paying rent. [When you are unable to pay rent] They take someone’s things or they close the house. – IDI2

As a result of these economic impacts, some people described being forced to move, either to their ancestral village if they had the means to get there, or to a cheaper, often smaller or less well-located, house within Nairobi’s slums:

> Life was very expensive, now it became very expensive to pay for rent… So, we had to find a cheaper life that we can sustain – IDI17

These indirect effects of the pandemic were gendered too. Male respondents frequently reported being especially responsible for earning money for the household, and females, including girls not attending closed schools, were more commonly responsible for childcare. However, there were also examples of where the crisis necessitated both parents to earn money for the household:

> I was able to provide but since I lost my job life became very expensive. I stayed in the house for long thinking of what to do. You have lost your job and you don’t have any other way, you don’t have anywhere to go. You don’t know how it is out there. Paying the house has become so hard you sometimes could stay for two or three months without paying rent. You have been given a notice… you don’t know where you are moving to. You see those are the challenges. You know that time [when I was employed] I didn’t have such challenges; I knew every month I have a salary and I also knew my family was catered for. I knew I am providing you see? So, when I lost my job, it became trial and error.-IDI17

### Impacts on young children span all domains of Nurturing Care

Secondly, the impacts of covid-19 controls on young children spanned all domains of nurturing care and were described in a variety of ways. Cutting across the domains, some parents/carers described the impacts of their own fear and a combination of confusion and some community denial about the epidemic, especially early on. For example, some reported how this led to them ‘shielding’ their children at home because of a combination of a strict interpretation of the restrictions and their own fear of the virus. This limited travel both within and beyond the city, reducing family and peer interaction, and leading to delaying or avoiding seeking of healthcare or shopping for food.

> People were fearing even to go to someone’s house or even greeting them… It happened that everyone was staying in the house and they don’t want to go out, you only run to the shop or fetch water and go back in the house – IDI7

Some parents/carers described how they were especially worried about their children’s risks from Covid-19 during the pandemic. This was either because of their desire to socialise with peers or because they were unable to use personal protective equipment that was thought to be effective like face masks:

> [I am afraid to travel] because she can’t put on a mask… So when I put on a mask to protect myself what of her? She will not allow me to cover her. She wants to look at everything – IDI12

### Health

Although no respondents reported Covid-19 making members of their household or extended household unwell, disruption to health services was a concern, for example with some child health clinics being converted into Covid-19 treatment or isolation centres leading to them not taking children, especially for health promotion and prevention interventions:

> But for now, [name of clinic] is for Corona patients… That is where we were taking a child for clinic – IDI12

Restrictions were described as placing considerable strain on all members of the household, including contributing to stress amongst parents/carers, with knock-on effects on the care of young children; discussed further below.

### Nutrition

In addition, significant knock-on effects of economic stress on food security was reported. Several study participants reported cutting down on meals to once a day, reducing the variety of food or relying on help from neighbours. The effects of this food insecurity were described as especially significant for children, accompanied by a sense of helplessness or lack of options. The challenge of managing on a day-to-day basis was clearly described:

> …We suffered, we stayed without. Sometimes we would take strong tea without sugar and the child will not drink … He would cry for the whole day but what can we do? You wake up in the morning you don’t have money and you find someone who gives you twenty shillings and you go and buy vegetables for ten shillings, a five-shilling tomato and an onion of five shillings and you add a lot of soup and you eat it – IDI5
>
> He would eat yoghurt and chips and all these things stopped … Now he just eats what has been found, strong tea… And you can see the sadness in his eyes … when he asks for something and you are unable to provide - IDI7

### Responsive Caregiving and Early Learning

Lockdown was described as mostly undermining both peer to peer and parent-child interactions, especially as restrictions became protracted. This included children being unable to play with their peers and becoming bored:

> Corona has affected them because they were playing as a group outside… So now you know he plays alone in the house so he is bored. This social distance thing. …he isn’t playing anymore – IDI9

As noted above, restrictions placed considerable strain on parents/carers mental wellbeing, and this context was described as having an impact on the parents’ ability to provide responsive care:

> I locked myself in the house and it reached a point and I said I better get sick with corona instead of seeing how children are crying daily… He would cry for the whole day but what can we do? – IDI5

Most childcare provision was reported to have closed when schools did. This was a result of several factors, including reduced demand because newly unemployed parents or older siblings (whose schools had closed) could now play a larger role in providing childcare at home, or because of parental concern about transmission risks in childcare:

> [Paid childcare] was not going on because you are fearing to take your child and meet with other children. Every parent was making the children fearful. – IDI7
>
> There were no jobs… we were not going [to paid childcare] when the schools were closed. All of us, even the children, were playing with him – IDI5

Parents/carers also reported cutting back on purchases of books, toys or clothes for the family, and being unable to afford school fees when schools reopened, in some cases leading to children being moved into cheaper schools when lockdown ended:

> It even became hard to pay for school. They were going to a good school so I had to transfer them – IDI21

When children were allowed to play, either alone, with family members, or with others in the community, this was also described as insufficient and leading to learning losses:

> You see during that time they were playing a lot… just playing. Playing is good but she was not reading at all. So, the things she had learnt, the teacher [at the pre-school] had to teach her again so she can catch up… she lagged a little. – IDI8

### Security and safety

When asked about levels and types of violence in their communities during this period, most study participants reported that Covid-19, and the ‘lockdowns’, led to an increase in the level of crime and domestic violence in their communities, or even in their own households. This worsening of community safety was described as being associated with economic hardship and food insecurity:

> [Domestic violence] is not far… even in my house. We are struggling a lot because of money… Because of money. One thing that makes people violent is money. Lack of money causes people to be violent… …when you have fifty shillings they see as if you have hidden another fifty shillings in your pocket. Such things, problems, is what makes people violent. Poverty. – IDI5
>
> There have been cases [of domestic violence]. You know, when people lack money mostly they disagree… So you can come and you were not successful to bring money for the day… you find they disagree and you fight. You know they are non-permanent houses – we hear people. Maybe they have disagreed because of money. Maybe one needs food and they haven’t provided. For that food they fight. – IDI17

### Help, when available, mostly came from within the community

One final major theme was that help, when provided, largely came from within the community. One example of this was informal sharing of limited food amongst neighbours:

> You visit the neighbour and eat the food they have. Or sometimes you hustle and go to borrow food like that. It may not be the immediate neighbour here you can go out and go on the other side and meet a friend – IDI7

In addition, flexibility in rent payments, including reductions or deferral of payments was important to many. Flexibility amongst landlords was described as being variable, with those who know and trust you being seen as more willing to help. There were also examples of community leaders (Chiefs) applying pressure to landlords to reduce the risk of evictions:

> Landlord has helped a lot because up until now we have not paid. But there is something that came and helped the people in iron sheets houses… Chief came and said people shouldn’t be pressured a lot about the houses – IDI5

Some respondents did talk about external assistance, including one who described benefiting from the government cash for work (Kazi kwa Vijana - Kenya Youth Empowerment, project), alongside intermittent food assistance, but this seemed to be uncommon and infrequent.

Overall, most experiences were relatively universal, with a consistent sense of the pandemic leading to a worsening of living conditions. However, and in contrast to most of the respondents, a small number of participants talked about how day to day life had in fact remained quite consistent. This was either because their own work hadn’t changed, or because life in the slums was very hard even before the pandemic. In addition, some described how they had managed to ‘shield’ their child from impacts of the pandemic:

> Because we are the one struggling so she can get what she needs … her life is going on as usual – IDI2

## Discussion

This research suggests that, despite being a low risk from SARS-CoV-2 infection, the covid-19 pandemic has radically affected the care of young children in Nairobi slums largely due to the direct and indirect effects of pandemic restrictions. These impacts are strikingly broad, affecting all domains of Nurturing Care, and deep, in terms of the scale of especially economic hardships.

### Key findings in context

Our findings are consistent with other published research yet provide additional child-centred insights. Considering the **cross-cutting** economic impacts, Oyando et al. (16) identified, through telephone surveys, high levels of economic and social disruption across three counties in Kenya, with especially pronounced effects on income, and amongst the poorest.

On **nutrition**, Kansiime et al. (17) looked specifically at food security impacts of Covid-19 across Kenya and Uganda, and found that more than two thirds (of a cohort completing an online survey) experienced income shocks and worsened food security, and Kimani-Murage et al. (18) concluded that restrictive covid-19 control measures exacerbated the pre-existing vulnerability amongst the urban poor to food insecurity and violated their human right to food; both consistent with the descriptions of widespread exacerbation of food insecurity amongst respondents in this study.

We found that covid-19 led to little reported direct harm to respondents, at least that they were aware of, but considerable disruption to **health** services. Oluich-Aridi et al. (19) conducted a qualitative study looking specifically at the impact of the pandemic on maternity services in Nairobi and identified many themes consistent with ours, including high levels of concern and perceptions of risk early in the pandemic, some reported reductions in access to maternal healthcare alongside significant economic harms including worsening food insecurity due to lockdowns and curfews. Ahmed et al. (20) also explored the impacts of covid-19 on access to healthcare across seven slums around the world, noting reduced access to services, increases in costs and fear discouraging utilisation.

A mixed-methods assessment of health effects of Covid-19 in Kenya found significant reductions in outpatient visits and – in keeping with the worsening community **safety** and increases in **domestic violence** reported in our study – an increase in sexual violence cases reported (21). The gendered aspects of the impacts extend beyond violence, however, as we found and as has also been noted by others in terms of the socio-economic impacts disproportionally affecting women and girls (22,23).

A recently published systematic review of the effects of Covid-19 on Nurturing Care around the world found an evidence base that was limited and biased towards high-income settings, and which suggested Covid-19 would lead to a need for increased support for young children to thrive in the pandemic(24). Particular priorities identified were the need to address parent/caregiver stress, burnout or depression and the potential for knock-on harsher parenting affecting **responsive caregiving and early learning**. The authors also identified a risk of reduced child safeguarding referrals and an urgent need for further research, including qualitative studies, to understand these risks in more depth.

Many of these priorities identified are consistent with our findings, for example regarding the disruption to childcare services and household tension at times affecting the care of young children. The absence of references to harsher parenting in our results probably reflects both the fact that we did not directly explore this issue and taboos around this subject.

As noted above, concerns were raised about the risks to young children by some before, during and after implementation of stringent covid-19 control measures in Kenya(5) and beyond (25,26). Yet, despite these warnings, our study suggests that only limited, and largely community drawn, support was received by families, suggesting that only limited policy attention and resources were devoted to these issues.

Overall, our findings suggest that concerns about the risks of ‘lockdown’ to young children in slums were largely well founded. Multiple harms or negative impacts of stringent covid-19 control measures on vulnerable young children growing up in slums were reported, and these spanned all domains of Nurturing Care.

### Strengths and limitations

There are a number of strengths to this study. Firstly, in-depth interviews allowed a deep exploration of parental perspectives and experiences during the pandemic. A purposively selected sample meant that a variety of parents/carers were interviewed, including male and female carers and those looking after different ages of children. This allowed gendered differences to be identified, although in general key themes were largely consistent across these groups. We were able to collect high quality data through interviews being conducted by an experienced researcher (RM) combined with regular analytical and reflexivity meetings.

A limitation of the research was, due to the prevailing covid-19 control measures, the use of an existing sampling frame which was based on prior, albeit recent, in-person enumeration of potential telephone survey respondents by our data collection partner. In addition, we were initially concerned that using remote data collection (telephone interviews) on occasions would present challenges to building rapport, although in practice telephone interviews worked better than we expected, with good rapport being built and few dropped calls.

### Unanswered questions and future research

The impacts reported by participants in this study ought to be explored in more detail, including through efforts to quantify their distribution and magnitude, and resultant impacts on child health and development. The NECS covid-19 impacts tracker has tracked disruption to early childhood services over time in Nairobi (manuscript in preparation), but studies from a variety of settings are urgently needed, including those that include measurement of child health and development outcomes.

Only through such research, alongside concurrent efforts to assess the real-life benefits of different Covid-19 control measures, can an informed discussion about the overall case for these types of pandemic control measures, in particular stringent ‘lockdowns’, be considered. Such analyses should inform the response to both future SARS-CoV-2 waves and other emergencies(25).

These results also imply an urgent need for both economic support and broader investment in public health and wellbeing for those living in slums, including in emergencies including epidemic disease outbreaks(27). Crucially, such investments are likely to be needed both in the short- and long-term to try to mitigate short term risks like food insecurity, and to ameliorate some of the longer-term harms including to early childhood development and education. In addition, longer-term investments in preparation for future crises are also needed(28).

## Conclusion

Based on the experiences of parents/carers, the Covid-19 pandemic, and efforts to control it, appear to have exacerbated adversity amongst young children growing up in slums in Nairobi. This includes through disrupting fragile and weak systems and placing considerable economic and social distress on vulnerable families and communities. Consideration of these insights can help to inform mitigation efforts and future epidemic control policy discussions. They imply that if blunt policy instruments like ‘lockdowns’ are to be used at all, then considerable efforts ought to be made to mitigate their associated harms, especially to young children growing up in informal settlements.

## Data Availability

We are happy to make available to all qualified researchers on request the underlying data drawn on for this research. Given the challenges of genuine anonymisation of qualitative data, we have not posted the data to an open repository, but we are happy to facilitate sharing requests via the corresponding author.

## Acknowledgements

We would like to acknowledge and thank the study participants for the time and insights that they shared with us to conduct this study, and to thank Antonio Aparicio at LSHTM and Pauline Ochieng at APRHC for vital administrative support to the study.

## Author contributions

RCH, BK, ZH & PK-W conceptualised the NECS study and RCH, PW & SB the linked covid-19 tracker study. RCH led the design and analysis for this paper and wrote the first draft. RM conducted the interviews and reviewed the draft manuscript, before discussing it with RCH and then all authors. RCH, RM, PK-W, SO and ZH regularly discussed and reviewed analytical coding as the data was collected. All authors edited this draft and provided important intellectual content. All authors approved the final manuscript.

## References

1. Aluga MA. Coronavirus Disease 2019 (COVID-19) in Kenya: Preparedness, response and transmissibility. J Microbiol Immunol Infect. 2020 Oct;53(5):671–3.

2. Bird J, Montebruno P, Regan T. Life in a slum: understanding living conditions in Nairobi’s slums across time and space. Oxf Rev Econ Policy. 2017 Jul 1;33(3):496–520.

3. Policing the Pandemic in Kenya – Policing the Lockdown [Internet]. [cited 2022 Sep 1]. Available from: https://blogs.ed.ac.uk/policingthelockdown-sipr/2020/05/08/policing-the-pandemic-in-kenya/

4. Black MM, Walker SP, Fernald LCH, Andersen CT, DiGirolamo AM, Lu C, et al. Early childhood development coming of age: science through the life course. The Lancet. 2017 Jan 7;389(10064):77–90.

5. Shumba C, Maina R, Mbuthia G, Kimani R, Mbugua S, Shah S, et al. Reorienting Nurturing Care for Early Childhood Development during the COVID-19 Pandemic in Kenya: A Review. International Journal of Environmental Research and Public Health. 2020 Jan;17(19):7028.

6. Hughes RC, Kitsao-Wekulo P, Bhopal S, Kimani-Murage EW, Hill Z, Kirkwood BR. Nairobi Early Childcare in Slums (NECS) Study Protocol: a mixed-methods exploration of paid early childcare in Mukuru slum, Nairobi. BMJ Paediatrics Open. 2020 Dec 1;4(1):e000822.

7. Kenya - COVID-19 Overview - Johns Hopkins [Internet]. Johns Hopkins Coronavirus Resource Center. [cited 2022 Aug 18]. Available from: https://coronavirus.jhu.edu/region/kenya

8. MINISTRY OF HEALTH – REPUBLIC OF KENYA [Internet]. [cited 2020 Jul 10]. Available from: https://www.health.go.ke/

9. The Busara Center for Behavioral Economics [Internet]. The Busara Center for Behavioral Economics. [cited 2020 Jul 10]. Available from: https://www.busaracenter.org

10. Chesire EJ, Orago AS, Oteba LP, Echoka E. Determinants Of Under Nutrition Among School Age Children In A Nairobi Peri-Urban Slum. East African Medical Journal. 2008;85(10):471–9.

11. Wamukoya M, Kadengye DT, Iddi S, Chikozho C. The Nairobi Urban Health and Demographic Surveillance of slum dwellers, 2002–2019:Value, processes, and challenges. Global Epidemiology. 2020 Nov 1;2:100024.

12. Hughes, R. C. et al. NECS COVID Impacts Tracker sub-study (NECS-cit) Protocol [Internet]. 2020. Available from: https://datacompass.lshtm.ac.uk/id/eprint/1780/

13. NVivo [cited 2022 Mar 12]. Available from: https://www.qsrinternational.com/nvivo-qualitative-data-analysis-software/home/

14. WHO | Nurturing care for early childhood development: Linking survive and thrive to transform health and human potential [Internet]. WHO. [cited 2019 May 17]. Available from: http://www.who.int/maternal_child_adolescent/child/nurturing-care-framework/en/

15. Kelly LM, Cordeiro M. Three principles of pragmatism for research on organizational processes. Methodological Innovations. 2020 May 1;13(2):2059799120937242.

16. Oyando R, Orangi S, Mwanga D, Pinchoff J, Abuya T, Muluve E, et al. Assessing equity and the determinants of socio-economic impacts of COVID-19: Results from a cross-sectional survey in three counties in Kenya [Internet]. Wellcome Open Research; 2021 [cited 2022 Aug 21]. Available from: https://wellcomeopenresearch.org/articles/6-339

17. Kansiime MK, Tambo JA, Mugambi I, Bundi M, Kara A, Owuor C. COVID-19 implications on household income and food security in Kenya and Uganda: Findings from a rapid assessment. World Development. 2021 Jan 1;137:105199.

18. COVID19 and Human Right To Food: Lived Experiences of the Urban Poor in Kenya with the Impacts of Government’s Response Measures, A Participatory Qualitative Study [Internet]. 2021 [cited 2022 Aug 31]. Available from: https://www.researchsquare.com

19. Oluoch-Aridi J, Chelagat T, Nyikuri MM, Onyango J, Guzman D, Makanga C, et al. COVID-19 Effect on Access to Maternal Health Services in Kenya. Frontiers in Global Women’s Health [Internet]. 2020 [cited 2022 Aug 21];1. Available from: https://www.frontiersin.org/articles/10.3389/fgwh.2020.599267

20. Ahmed SAKS, Ajisola M, Azeem K, Bakibinga P, Chen YF, Choudhury NN, et al. Impact of the societal response to COVID-19 on access to healthcare for non-COVID-19 health issues in slum communities of Bangladesh, Kenya, Nigeria and Pakistan: results of pre-COVID and COVID-19 lockdown stakeholder engagements. BMJ Global Health. 2020 Aug 1;5(8):e003042.

21. Barasa E, Kazungu J, Orangi S, Kabia E, Ogero M, Kasera K. Indirect health effects of the COVID-19 pandemic in Kenya: a mixed methods assessment. BMC Health Services Research. 2021 Jul 26;21(1):740.

22. Kithiia J, Wanyonyi I, Maina J, Jefwa T, Gamoyo M. The socio-economic impacts of Covid-19 restrictions: Data from the coastal city of Mombasa, Kenya. Data in Brief. 2020 Dec 1;33:106317.

23. Decker MR, Wood SN, Thiongo M, Byrne ME, Devoto B, Morgan R, et al. Gendered health, economic, social and safety impact of COVID-19 on adolescents and young adults in Nairobi, Kenya. PLOS ONE. 2021 Nov 9;16(11):e0259583.

24. Proulx K, Lenzi-Weisbecker R, Hatch R, Hackett K, Omoeva C, Cavallera V, et al. Nurturing care during COVID-19: a rapid review of early evidence. BMJ Open. 2022 Jun 1;12(6):e050417.

25. Innocenti UO of R. What were the immediate effects of life in lockdown on children? [Internet]. UNICEF-IRC. [cited 2022 Mar 14]. Available from: https://www.unicef-irc.org/article/2163-what-were-the-immediate-effects-of-life-in-lockdown-on-children.html

26. UNSDG | Policy Brief: The Impact of COVID-19 on children [Internet]. [cited 2022 Mar 12]. Available from: https://unsdg.un.org/resources/policy-brief-impact-covid-19-children, https://unsdg.un.org/resources/policy-brief-impact-covid-19-children

27. Lilford RJ, Oyebode O, Satterthwaite D, Melendez-Torres GJ, Chen YF, Mberu B, et al. Improving the health and welfare of people who live in slums. The Lancet. 2017 Feb 4;389(10068):559–70.

28. Main Report & accompanying work [Internet]. The Independent Panel for Pandemic Preparedness and Response. [cited 2022 Aug 21]. Available from: https://theindependentpanel.org/mainreport/

